# The effect of notification window length on the epidemiological impact of COVID-19 contact tracing mobile applications

**DOI:** 10.1101/2021.11.08.21266079

**Authors:** Trystan Leng, Edward M. Hill, Matt J. Keeling, Michael J. Tildesley, Robin N. Thompson

**Affiliations:** The Zeeman Institute for Systems Biology & Infectious Disease Epidemiology Research, School of Life Sciences and Mathematics Institute, University of Warwick, Coventry, CV4 7AL, United Kingdom; JUNIPER – Joint UNIversities Pandemic and Epidemiological Research, https://maths.org/juniper/

## Abstract

The reduction in SARS-CoV-2 transmission from contact tracing applications (apps) depends both on the number of contacts notified and on the probability that those contacts quarantine after notification. Referring to the number of days preceding a positive test that contacts are notified as an app’s *notification window*, we use an epidemiological model of SARS-CoV-2 transmission that captures the profile of infection to consider the trade-off between notification window length and active app-usage. We focus on 5-day and 2-day windows, the lengths used by the NHS COVID-19 app in England and Wales before and after 2nd August 2021, respectively. Short windows can be more effective at reducing transmission if they are associated with higher levels of active app usage and adherence to isolation upon notification, demonstrating the importance of understanding adherence to control measures when setting notification windows for COVID-19 apps.

## 2 Main

The automated tracing of close contacts via mobile phone applications (apps) has been used in many countries to reduce transmission of SARS-CoV-2^1^. In England and Wales, the National Health Service (NHS) COVID-19 contact tracing app has been available since 24th September 2020^2^. After a user submits a positive test result, the app identifies via bluetooth their recent high-risk encounters with other app users, with a high-risk encounter defined as being within two metres of someone for at least fifteen minutes^3^. If the user is symptomatic upon submitting a test, high-risk encounters *d* days prior to symptom onset up until the present moment are identified, while if a user is asymptomatic, high-risk encounters *d* days prior to the individual taking a test up until the present moment are identified. Contacts identified as involved in a high-risk encounter are then notified of potential exposure. We refer to *d* as an app’s *notification window*. An app’s effectiveness at reducing transmission depends on the number of contacts who are identified through an app. It might therefore be expected that longer notification windows will lead to greater reductions in transmission. However, long notification windows may have negative consequences, such as a large number of notifications being issued to uninfected individuals, with potential impacts on app usage. In England, over one million notifications were sent to contacts in the first two weeks of July 2021^4^, leading some commentators to suggest that many individuals identified through the app were isolating unnecessarily. On 2nd August 2021 the notification window was reduced from 5 days to 2 days for asymptomatic individuals submitting a positive result^5^, to encourage the continued use of the NHS COVID-19 app while limiting the number of individuals isolating.

If further measures are necessary to mitigate SARS-CoV-2 transmission in the future, increasing the notification window would seem an intuitive response because the number of potential infectious contacts notified would increase. However, the effectiveness of contact tracing not only depends on the number of infected individuals notified, but also depends on people’s behaviour upon notification^6^. In part, an app’s effectiveness depends upon the likelihood that an individual’s contacts actively use the app and adhere to the recommended self-isolation period, which in turn depends on the perceived risk that a notified individual is infected. As well as increasing the number of infectious contacts notified, longer notification windows increase the number of uninfected individuals asked to self-isolate, which may reduce public confidence lead to lower levels of active app use. Here, we analyse the expected number of primary cases that result from a base case who reports their infection to a contact tracing app. We then consider the expected number of secondary cases that result from those primary cases (illustrated in Figure 1), and explore the effectiveness of app-based notifications at reducing transmission with either 2-day and 5-day notification windows at different levels of active app use. Rather than aiming to generate precise quantitative predictions, our goal is to use a simple epidemiological model to explore the general impacts of different notification windows on SARS-CoV-2 transmission.

**Figure 1:**
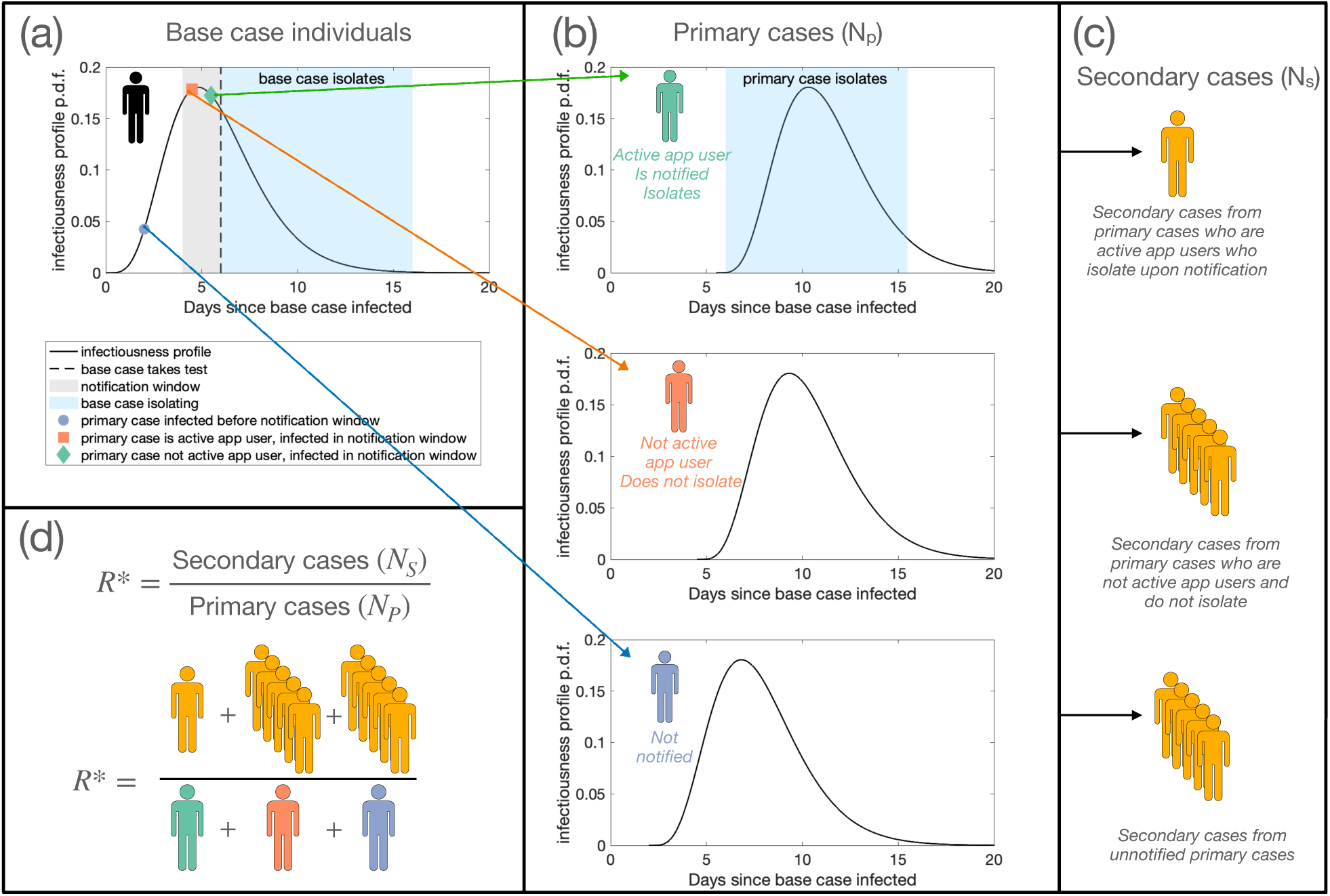
Schematic of the modelling approach. (a) The infectiousness of a base case individual varies through time. A base case individual takes a test on day *d* in their infectiousness profile (dashed line), after which they isolate (light blue shaded area). (b) Primary cases who have the app that are infected by the base case from day *d− w* to day *d* are notified of possible exposure. A proportion *p* of individuals infected within the notification window adhere to self-isolation (green, top panel), while (1*− p*) of individuals are not active app users, and do not self-isolate (orange, middle panel). Adhering individuals self-isolate from notification until *i*_*notif*_ days have elapsed since contact with the base case. Primary cases infected before day *d− w* are not notified (blue, bottom panel). Those who do not adhere to isolation upon notification or who are not notified either remain mixing with the population with either no interventions placed upon them throughout their infectious period or they isolate upon onset of symptoms. (c) The expected number of secondary cases from non-adhering or unnotified primary cases is higher than the expected number of secondary cases that result from primary cases who adhere to isolation after notification. (d) *R*^∗^ is calculated as the ratio between the expected number of secondary cases and the expected number of primary cases - in our illustrative example, the expected number of secondary cases is 11, and the expected number of primary cases is 3, giving *R*^∗^ = 11/3.

Using our epidemiological model, we consider a base case app user who becomes infected on day 0 and who tests positive to SARS-CoV-2 on day *d*. To explore the effect of notification window length on transmission in a concrete setting, we consider an asymptomatic base case who is detected using a lateral flow device test (LFT), with no delay between taking a test and receiving a result (for an instance where a base case seeks a PCR test, with a two day delay, see Supplement S2). The relative probability of a base case testing positive on a given day *d* varies through time (Figure 2a), which we obtained by normalising a previously published test probability profile for LFTs^7^. This is equivalent to assuming that the base case tests once at a random time during infection, and is detected - the impact of more regular testing is considered in Supplement S3. We assume that the base case self-isolates after taking a test. The expected number of primary infections from the base case prior to taking a test is informed by a previously derived SARS-CoV-2 infectiousness profile^8^, under the assumption that contacts occur randomly at a constant rate until taking a test. Primary cases infected within the notification window (grey shaded area in Figure 1a) are notified of possible exposure, while those infected before the notification window receive no notification. Primary cases infected within the notification window self-isolate with probability *p*, with *p* representing the proportion of the population who are active app users. We define active app use as both having the app (downloaded and with bluetooth enabled) and adhering to isolation upon notification. Those who are infected before the notification window or are not active app users continue mixing with the population throughout their infectious period if asymptomatic, or if symptomatic until symptom onset, at which point they self-isolate. We assume that asymptomatic cases comprise 30% of all cases^9^ and are 50% as infectious as symptomatic cases^10^. We use a previously derived distribution of incubation periods^11^ to determine the time from infection to symptom onset for symptomatic cases. To estimate the number of infections that each primary case is expected to generate, we directly calculate the effective reproduction number, *R*^∗^, as the ratio between the expected number of secondary infections and the expected number of primary infections (Figure 1d). See Supplement S1 for a detailed explanation of the methods.

**Figure 2:**
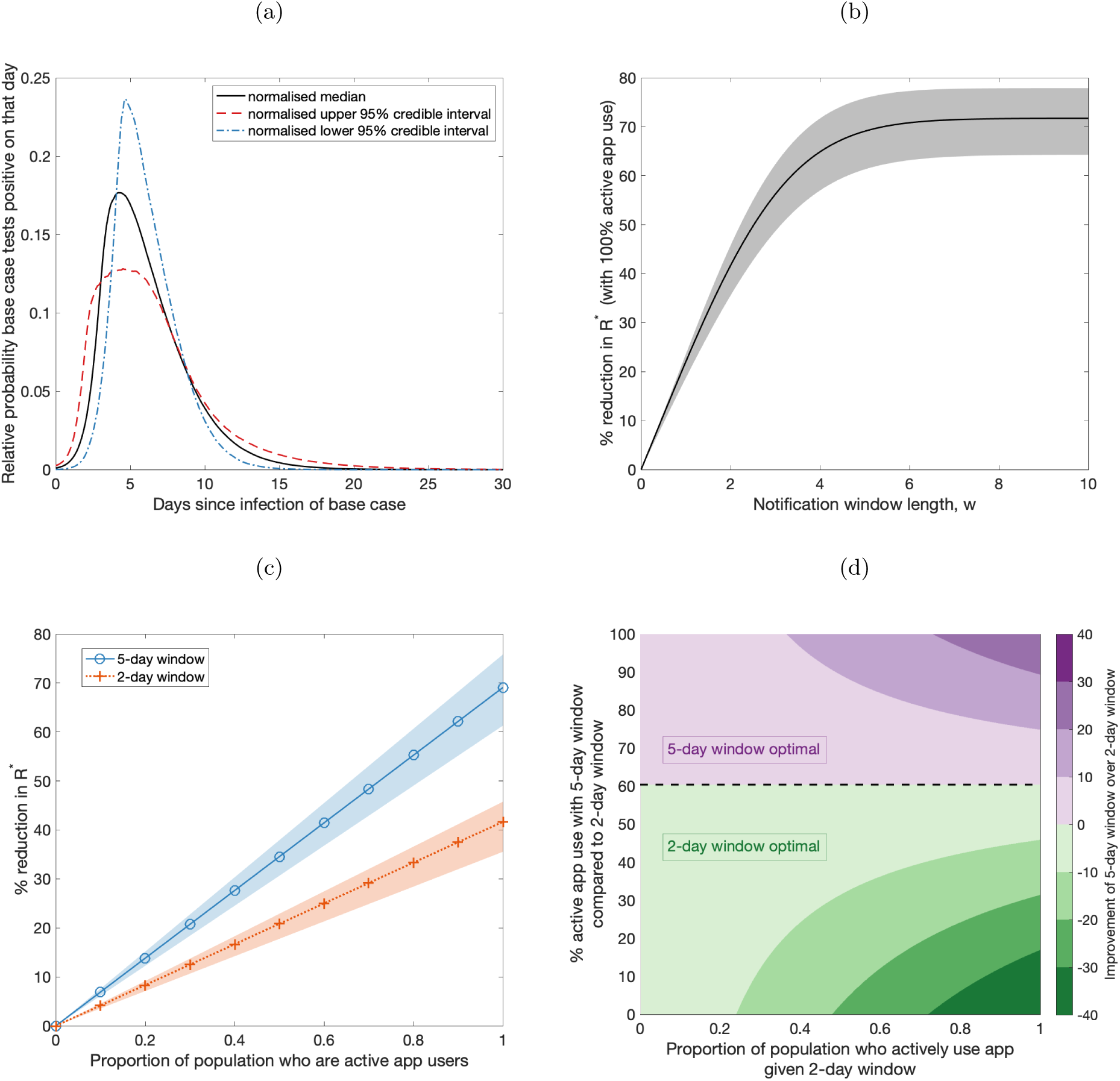
The impact of notification window length and adherence on transmission. (a) The relative probability of the base case testing positive on a given day in their infectiousness profile, obtained by normalising the median (black, solid line) test probability profiles of LFTs for asymptomatic testing^7^. Normalised 95% credible interval test probability profiles^7^ (upper - red, dashed line; lower - blue, dot-dashed) are also used, to obtain shaded regions in (b) and (c). (b) The percentage reduction in *R*^∗^ for different length notification window *w*, relative to a scenario in which a notification app is not used, under the assumption that all notified individuals adhere to isolation upon notification. (c) The relationship between the proportion of primary cases who are active app users and percentage reduction in *R*^∗^ for 5-day notification windows (blue solid line, circle markers) and 2-day notification windows (orange dotted line, cross markers). (d) A heat map of the improvement of a 5-day window over a 2-day window, quantified by the difference in the percentage reduction in *R*^∗^ that results from a 5-day notification window compared to a 2-day notification window. The proportion of primary cases assumed to be active app users for a 2-day window is shown on the x-axis, and the relative level of active app use assumed for a 5-day window (compared to the level of active app use for a 2-day window) is shown on the y-axis. Purple (green) regions correspond to where 5-day (2-day) notification windows lead to a larger reduction in *R*^∗^.

We first consider the impact of notification window length on transmission assuming all primary cases are active app users, i.e. all primary cases infected within the notification window adhere to self-isolation upon notification. The reduction in *R*^∗^ resulting from app-based notifications (relative to a scenario in which a notification app is not used) increases with the notification window length (Figure 2b), though there are only limited further benefits in transmission reduction for notification windows longer than 5 days. Assuming 100% active app use, a 2-day notification window results in a 42% reduction in *R*^∗^ while a 5-day notification window results in a 69% reduction in *R*^∗^. Even long notification windows do not eliminate transmission entirely, as primary cases may transmit infection before they are notified of their contact with the base case.

Next, considering more realistic scenarios where not all individuals are active app users, we calculate the reduction in *R*^∗^ for different levels of active app use. For a given notification window duration, the reduction in *R*^∗^ increases linearly with the proportion of primary cases who actively use the app and adhere to isolation (Figure 2c). A higher level of active app use is required to obtain a given reduction in *R*^∗^ for a 2-day window than for a 5-day window. For example, with a 5-day window, 58% of primary cases must be active app users to reduce *R*^∗^ by 40%, while a 2-day window requires 96% of primary cases to be active app users to obtain the same 40% reduction in *R*^∗^. If the proportion of the population who actively use the app is high (above 60%), then a 5-day window is always more effective than a 2-day window, irrespective of the level of active app use with a 2-day window.

Assuming the same level of active app use, longer notification windows result in larger reductions in transmission. However, if fewer individuals are active app users when a longer notification window is chosen, then a 2-day window can lead to a greater reduction in transmission than a 5-day window (Figure 2d). Under the model considered here, when the level of active app use given a 5-day window is less than 60% of the level of active app use given a 2-day window, the 2-day window results in less transmission. In other words, if more than 4 out of 10 individuals who would actively use the app under a 2-day window would no longer actively use the app under a 5-day window, then the increase in secondary cases from individuals no longer actively engaging with the app is greater than the reduction in secondary cases that results from capturing more primary cases in the notification window.

In our main analysis, we assume that base cases test positive to an LFT on a random day in their infectious period. In reality, this detection time distribution has varied throughout the course of the epidemic^12^ and will be influenced by a variety of factors, such as the requirements of different workplaces regarding regular testing. Understanding this distribution is important, as the relative impact of different notification windows at different levels of active app use depends upon when base cases are detected (Supplement S3). Early detection not only reduces the expected number of secondary cases from infected individuals, but also increases the proportion of secondary cases captured by shorter notification windows. If cases are detected early, the secondary cases missed by shorter windows may be offset by modest increases in active app use.

As with any epidemiological modelling study, our analyses involve a number of simplifying assumptions. We do not explicitly consider the impact of vaccination, which may impact results in two opposite directions. On the one hand, as vaccinated individuals are less likely to become infected^13^ and transmit the virus^14^, their inclusion would be expected to reduce transmission. On the other, in England fully vaccinated individuals have not been legally required to self-isolate upon notification since 16th August 2021^15^, increasing the number of high-risk encounters vaccinated individuals may have. These opposing factors make the impact of vaccination on overall app effectiveness unclear. In our model, we assume contacts occur at random and are not repeated, so a 2-day window will find 40% of the contacts a 5-day window would find. In reality, this proportion would be expected to be higher because of repeat contacts. We also assume that the infectiousness profile of symptomatic and asymptomatic individuals are the same, and that the infectiousness profile of symptomatic individuals and their incubation period are independent functions of time. This simplifying assumption is common in the literature^16,17^, although methods relaxing this assumption have also been explored^8,18^, providing an avenue for future research.

Adopting a parsimonious approach, our results demonstrate the complexity in assessing the optimal length of notification windows for mobile contact-tracing apps. A 5-day window is considerably more effective at reducing transmission than a 2-day window if the level of active app use in the population is not affected by the notification window length. If high levels of active app use can be achieved for a 5-day window, this strategy will always be optimal. However, as of 27th October 2021, there had been approximately 28.6 million downloads of the NHS COVID-19 app in England and Wales^4^. With an adult population of over 48 million^19^, a substantial proportion of eligible users must not have the app. Moreover, results from a large study of adherence to self-isolation in the UK^20^ from March 2020 to January 2021 indicate relatively low adherence to self-isolation measures after notification via contact tracing; while 65% of respondents reported an intention to adhere to self-isolation after being contacted, only 11% of contacted individuals did not leave home for 14 days after being contacted (the self-isolation policy implemented during the study period). If levels of active app use are low, and if individuals are less likely to actively use the app for longer notification windows, then the benefits of a longer notification window are outweighed by the costs of lower active app use. When making decisions about the length of notification window, policy makers should consider the potential impacts on active app use, and the resulting effects on transmission. We have provided a framework for guiding these decisions.

## Contributors

R.T. and M.J.T. conceptualised and designed the methodology of this study. T.L. wrote the model code, undertook the modelling analysis, and analysed and visualised the results. E.M.H. validated the model code and results. T.L wrote the original draft of the manuscript, and all authors reviewed and edited the manuscript.

## Acknowledgements

We thank Christopher Davis and Laura Guzmán Rincón for comments on an early draft, and Laura Guzmán Rincón for reviewing the model code.

TL, MJK and MJT were supported by UKRI through the JUNIPER modelling consortium [grant number MR/V038613/1]. MJK, RNT and MJT were supported by the Engineering and Physical Sciences Research Council through the MathSys CDT [grant number EP/S022244/1]. EMH, MJK and MJT were supported by the Biotechnology and Biological Sciences Research Council [grant number: BB/S01750X/1]. RNT was supported by UKRI through the Rapid Assistance in Modelling the Pandemic continuity grant [grant number: EP/V053507/1]. MJK was supported by the National Institute for Health Research (NIHR) [Policy Research Programme, Mathematical & Economic Modelling for Vaccination and Immunisation Evaluation, and Emergency Response; NIHR200411]. MJK is affiliated to the National Institute for Health Research Health Protection Research Unit (NIHR HPRU) in Gastrointestinal Infections at University of Liverpool in partnership with UK Health Security Agency (UKHSA), in collaboration with University of Warwick. MJK is also affiliated to the National Institute for Health Research Health Protection Research Unit (NIHR HPRU) in Genomics and Enabling Data at University of Warwick in partnership with UK Health Security Agency (UKHSA). The views expressed are those of the author(s) and not necessarily those of the NHS, the NIHR, the Department of Health and Social Care or UK Health Security Agency. The funders had no role in study design, data collection and analysis, decision to publish, or preparation of the manuscript.

## Declaration of interests

We declare no competing interests.

## Data availability

Data used to parameterise the study is publicly available and stated within the main manuscript and Supporting Information. Code for the study is available at: https://github.com/tsleng93/AppModelling

## Supplementary Materials

### S1 Methods

In this analysis, we consider a base case asymptomatic app user infected on day 0 who tests positive and self-isolates from day *d*. The relative probability a base individual is detected on day *d* depends on the likelihood that they test positive upon taking a test. We define *T*(*d*) as the probability density function of the time in which a base case is detected. We obtain this using a test probability profile for lateral flow tests for asymptomatic testing^7^, assuming that detected cases are sampled according to their probability of testing positive that day. By considering the expected number of primary cases that are infected by a base case individual, the expected number of secondary cases that result from those primary cases, and by varying the probability *p* that notified primary cases are active app users and adhere to their notification and consequently self-isolate, we can understand the reduction in transmission that results from different levels of active app use for different notification windows.

The expected number of infections resulting from an infected individual depends on an individual’s infectivity profile through time since becoming infected, as well as on the time periods when an individual is not isolating. We make the simplifying assumption that, assuming an individual is not isolating, their contact rate is constant through time. We let *R* denote the expected number of secondary infections from a symptomatic individual, assuming that a symptomatic individual mixes within the population with no interventions placed upon them for the entirety of their infectious period. Letting *f*(*t*) be the probability density function of an infected individual’s infectivity profile through time, we set *f* = Γ(5.62, 0.98), corresponding to a previously derived infectivity profile obtained using data from known infector–infectee pairs^8^. We let *F*(*d*) represent the cumulative density function of the infectivity profile i.e. 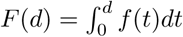.

Assuming the base case individual tests positive and inputs their result to their app, all contacts of the base case from time *d−w* to *d* are notified of possible exposure, where *w* denotes the notification window length (in days). Our analysis focuses on the cases *w* = 5 and *w* = 2, the notification window lengths used in the UK prior to and after the 2nd August 2021 respectively^5^. Adhering notified individuals isolate for *i*_*notif*_ days upon being notified, counted from the time of last contact. We assume that the time of last contact between the base case and primary case individual was the time the primary case individual was infected by the base case. Notified individuals are expected to isolate for 10 **full** days from the day of last contact^15^ - accordingly, we set *i*_*notif*_ = 10.5.

We let *l* denote the delay between the base case taking a test and receiving a result. We assume the base case takes a lateral flow test (LFT) and set *l* = 0. In Supplement S2, we consider an instance where the base case individual takes a PCR test, assuming *l* = 2.

The period an infected individual isolates for will depend on whether they develop symptoms. We assume that a proportion *A* of primary cases are asymptomatic. Here we set *A* = 0.3, in line with estimates of the proportion of SARS-CoV-2 infections that are asymptomatic reported in a meta-analysis^9^. We assume the same infectivity profile for symptomatic and asymptomatic individuals, but assume that asymptomatic individuals are relatively less infectious by a factor *α*, 0 < *α* ≤ 1. We used *α* = 0.5, in line with previous modelling studies, though estimates obtained from empirical studies vary substantially^10^. Upon symptom onset, we assume that individuals test positive and isolate for a period of *i*_*sympt*_ days. In the UK, symptomatic individuals are expected to isolate for 10 full days from symptom onset - accordingly, we set *i*_*sympt*_ = 10.5^21^. Letting *s*(*t*) be the probability density function of the time to symptom onset from day of infection for symptomatic individuals, we set *s*(*t*) = Γ(5.807, 0.9493)^22^. We make the assumption that *s*(*t*) and *f*(*t*) are independent functions of time - while this is a simplifying assumption, it is one common in the literature^16,17^.

A proportion *p* of the primary cases are assumed to be active app users. Active app users are defined as individuals who both a) have and use the app and b) adhere to isolation upon notification. In our analyses we vary *p* from 0 to 1.

Below, we consider the expected number of primary cases and secondary cases in turn.

### S1.1 Primary cases

If a base case takes a test on day *d* and isolates accordingly from day *d*, then the expected number of infections from the base case prior to isolation is given by:

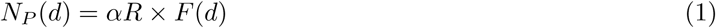

The expected number of infections from a typical base case prior to isolation is given by integrating over the day a base case takes a test, and accounting for the relative probability an individual tests positive that day:

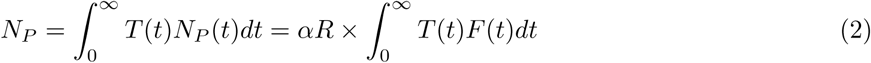

### S1.2 Secondary cases

Secondary cases infected by primary cases can occur in three different ways:

1. **Secondary cases resulting from primary cases infected before the notification window** All primary cases infected by the base case from day 0 to day *d* − *w* are not notified of exposure. Asymptomatic individuals remain infectious throughout their infectious period, while symptomatic individuals remain infectious until symptom onset before isolating for *i*_*sympt*_ days. The expected number of secondary infections in this instance is given by:

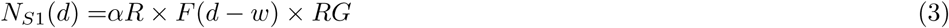

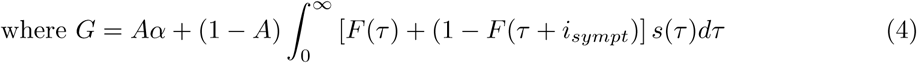

The expected number of secondary infections in this instance from a typical base case is given by:

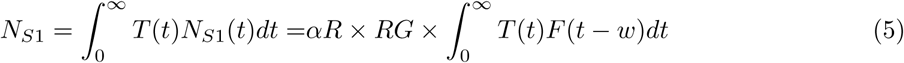
2. **Secondary cases resulting from primary cases infected within the notification window, but are not active app users** Primary cases infected from day *d*−*w* to day *d* are infected within the notification window. A proportion (1*− p*) of such cases are not active app users, and therefore either do not receive a notification, or do not adhere to isolation upon notification. Asymptomatic individuals who are not active app users remain infectious throughout their infectious period, while symptomatic individuals who are not active app users remain infectious until symptom onset, then isolate for *i*_*sympt*_ days. Hence, the expected number of secondary infections in this instance is given by:

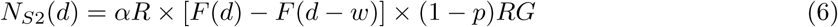

The expected number of secondary infections in this instance from a typical base case is given by:

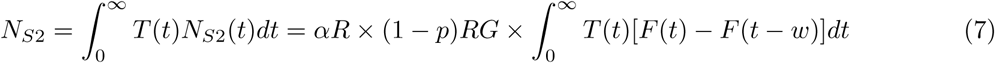
3. **Secondary cases resulting from primary cases infected within the notification window who are active app users** A proportion *p* of primary cases infected from day *d*− *w* to *d* are active app users. These primary cases are notified of possible exposure and adhere to isolation upon notification. They remain infectious until day *d* + *l*, and could be anywhere from day *l* to day *w* + *l* of their infectious period. In this case, there are four subcases: *Subcase a* (*N*_*S*3*a*_(*d*)): Asymptomatic individuals isolate from day *d* + *l*, and stop isolating *i*_*notif*_ days from their last day of contact with the base case.

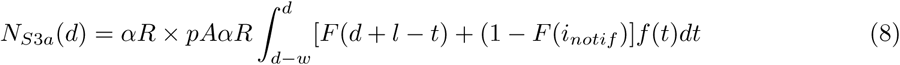

*Subcase b* (*N*_*S*3*b*_(*d*)): Symptomatic individuals who exhibit symptoms before day *d* + *l*, i.e. before their notification. These individuals are assumed to isolate for *i*_*sympt*_ days from symptom onset, with this isolation period not extended by their subsequent close contact notification.

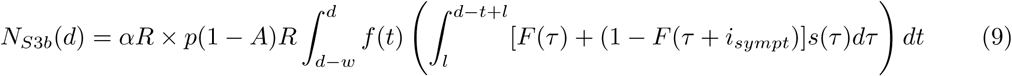

*Subcase c* (*N*_*S*3*c*_(*d*)): Symptomatic individuals infected on day *t* who exhibit symptoms after day *d* + *l* but before day *d* + *i*_*notif*_ − *t*, i.e. while they are isolating from their notification. These individuals isolate from *d* + *l* until *i*_*sympt*_ days after the onset of their symptoms.

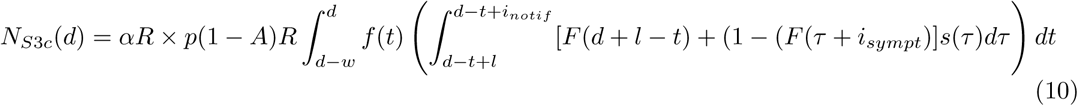

*Subcase d* (*N*_*S*3*d*_(*d*)): Symptomatic individuals infected on day *t* who exhibit symptoms after *d* + *i t*, i.e. after they have finished isolation from their notification. These individuals isolate from *d* + *l* until *d* + *i t*, and then leave isolation before isolating once again upon symptom onset, for *i*_*sympt*_ days from symptom onset.

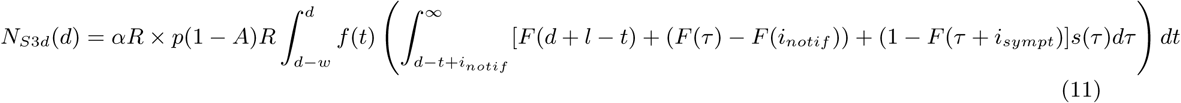

Together, we obtain

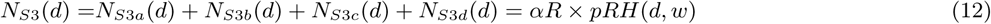

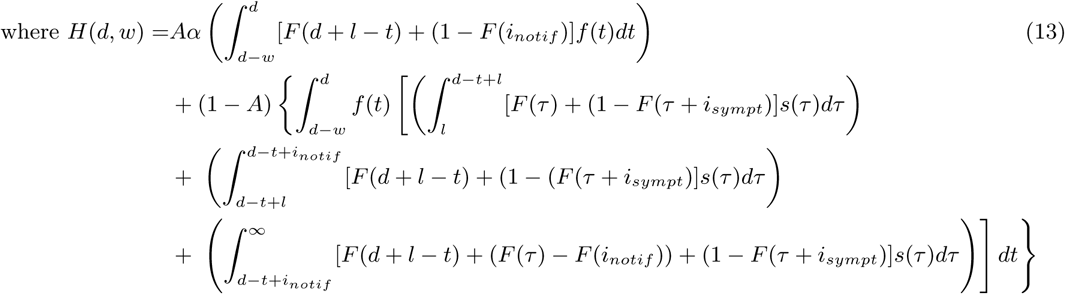

The expected number of secondary infections in this instance from a typical base case is given by:

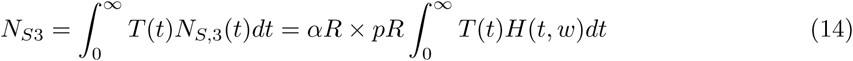

### S1.3 Calculating effective R

The effective reproduction number, *R*^∗^ is calculated as the ratio between the expected number of primary cases and the expected number of secondary cases:

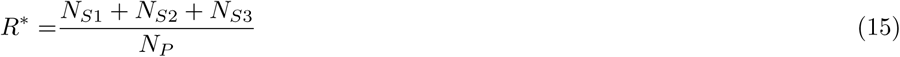

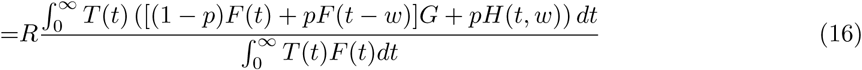

### S1.4 Determining increase in adherence required for a 2-day window to be optimal

Let *p*_*w*_ denote the proportion of the population who are active users that the app implements a *w*-day window, and let *R*^∗^(*w*) denote the effective reproduction number given a notification window of *w*. Letting *p*_5_ = *ϵp*_2_, 0 ≤ *ϵ* ≤ 1, we observe that the value of *ϵ* required for a 5-day window to be optimal is independent of *p*_2_:

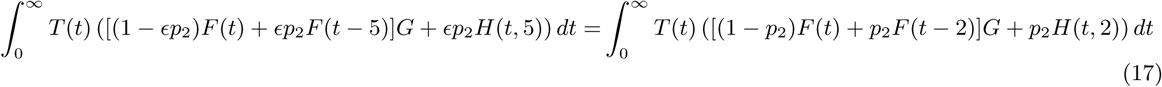

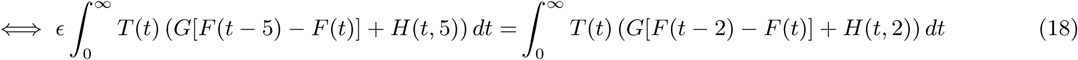

For a given window *w, R*^∗^(*w*) decreases as *p*_*w*_ increases if and only if 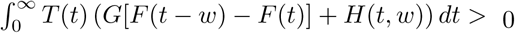. Otherwise, *R*^∗^(*w*) **increases** with increasing active app use. These instances are not considered here, as the intended control measure increases transmission. Given that *R*^∗^(*w*) decreases as active app use increases, a 2-day window is optimal given that *ϵ* satisfies:

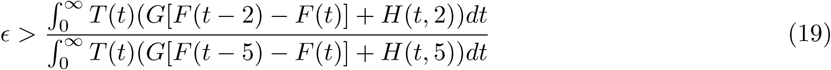

### S2 Assuming the base case seeks a PCR test

This section considers a scenario where base cases are detected through a PCR test, rather than through an LFT. While LFTs can produce a result in 30 minutes, PCR tests must be processed in a laboratory. Accordingly, there is a delay in individuals receiving a result, with consequential delays in their potential infectious contacts being notified. Here, we assume a delay of 2 days. In Figure S1, we observe qualitatively similar results regarding the impact of notification window length and active app use to those obtained assuming no delay. However, because of the delay in notification of primary cases, the transmission reduction from app-based notifications is considerably lower, with 2-day and 5-day notification windows resulting in a 17% and 25% reduction in transmission assuming an entire population of active app users, respectively.

### S3 Exploring the impact of the day of base case detection

Assuming that not all individuals are active app users, the reduction in transmission from the app depends upon: the day in which a base case individual is detected, *d*; on the number of their contacts notified (determined by the length of notification window, *w*); and on the proportion *p* of the population who are active app users, who adhere to self-isolation upon notification (Figures S2a and S2b). For *p <* 0.1, neither of the considered notification window lengths reduces *R*^∗^ by more than 10%. There is no difference in the reduction in *R*^∗^ when *d <* 2 because all infectious contacts are captured by both notification windows. However, a 2-day notification window can be considerably less effective when base case individuals are detected at a later stage of their infection (Figure S2c). The greatest difference in *R*^∗^ occurs when the base case is detected between 7 and 8 days into their infectiousness profile, with the difference in *R*^∗^ increasing with probability of adherence.

However, assuming that fewer individuals are likely to be active app users for a 5-day window than for a 2-day window, a 2-day window can become optimal for some regions of (*d, p*) space (Figures S2d to S2f). Assuming that active app use is 80%, 60%, and 40% of the active app use given a 2-day notification window, a 2-day notification window is optimal when *d <* 4.8 days, *d <* 6.1 days and *d <* 7.9 days respectively, with the greatest differences occurring when assuming a high level of active app use and very early detection of the base case individual.

Assuming the base case individual tests positive on day *d* from a one-off test (Figure S3a), and assuming a proportion *p* of primary cases are active app users, the relative level of active app use required given a 5-day window to be optimal is independent of *p* (Figure S3b, solid black line). If the base case tests positive early in their infectiousness profile, high relative levels of active app use are required for a 5-day notification window to beneficial, while if a base case is detected late, a 5-day window may remain optimal despite the lower level of active app use. Because of this, it becomes increasingly important to identify asymptomatic cases early in their infection profiles for shorter notification windows. In our main analyses, we consider a base case individual who tests positive on a random day in their infectiousness profile. Here, we consider an alternative situation where the base case individual takes regular tests every *r* days. The probability an individual tests positive on a given day *d* when *d < r* can be obtained directly from a test probability profile, while the probability an individual tests positive on day *d* when *d > r* is the probability that individual tests positive on day *d* and does not test positive on subsequent test days, which can be inferred from a test probability profile. Individuals are more likely to be identified early if they engage in regular mass testing (Figure S3a). Consequently, the required level of active app use given a 5-day window for that strategy to remain optimal increases. As an example, assuming a base case takes a lateral flow test every seven days, a 5-day window is optimal given that active app use is at least 62% of the level of active app use given a 2-day window (Figure S3b, purple square); assuming a base case takes a lateral flow test every three days, a 5-day window is optimal given that active app use is at least 70% of the level of active app use given a 2-day window (Figure S3b, pink diamond).

**Figure S1:**
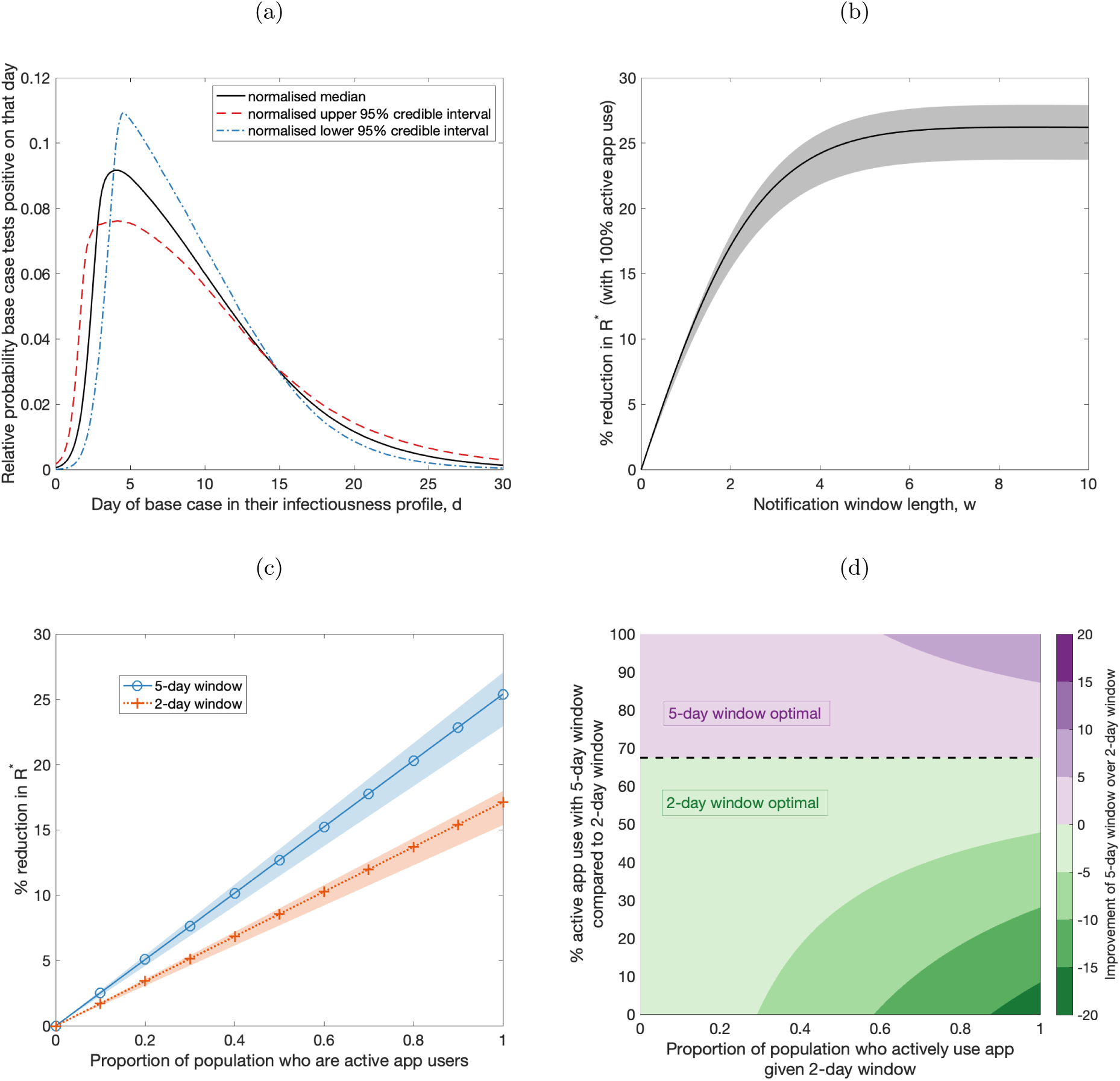
The impact of notification window length and adherence on transmission reduction, assuming the base case is detected through a PCR test. (a) The relative probability of the base case testing positive on a given day in their infectiousness profile, obtained by normalising the median (black, solid line) test probability profiles of PCR tests for asymptomatic testing^7^. Normalised 95% credible interval test probability profiles (upper - red, dashed line; lower - blue, dot-dashed) are used to obtain shaded regions for (b) and (c). (b) The percentage reduction in *R*^∗^ with respect to increasing length of the notification window *w*, under the assumption that all notified individuals adhere to isolation upon notification. (c) The relationship between probability of adherence and percentage reduction in *R*^∗^ for and 5-day notification windows (blue solid line, circle markers) and 2-day notification windows (orange dotted line, cross markers). A heat map of the improvement of a 5-day window over a 2-day window, quantified by the difference in the percentage reduction in *R*^∗^ that results from a 5-day and 2-day notification window, with the probability of adherence given a 2-day window on the x-axis, and the percentage reduction in adherence given a 5-day window compared to the probability of adherence given a 2-day window on the y-axis. Purple (green) regions correspond to where 5-day (2-day) notification windows lead to a larger reduction in *R*^∗^.

**Figure S2:**
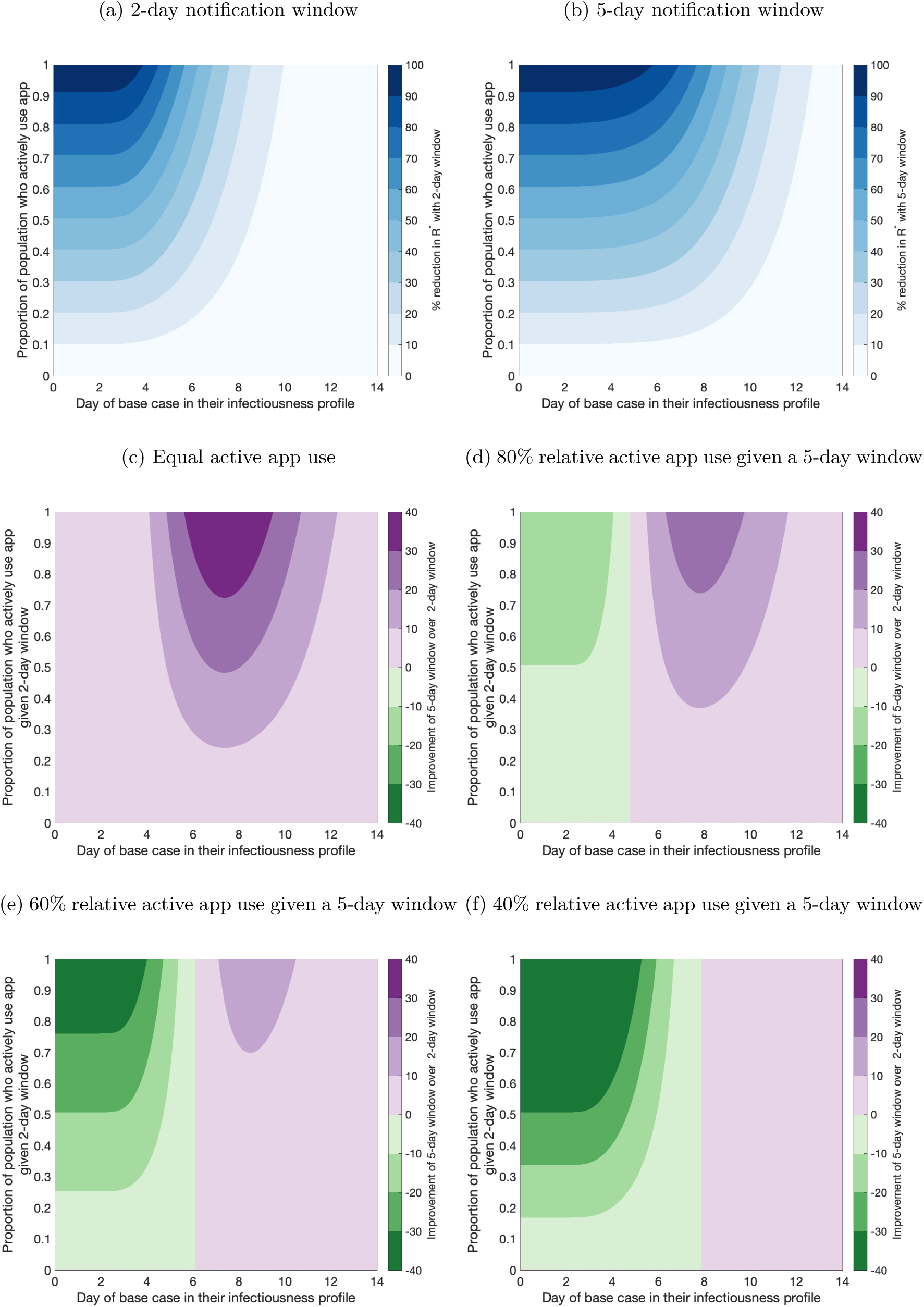
The impact of detection time of base case and active app use on transmission reduction. (a & b) The percentage reduction in *R*^∗^ that results from a 2-day and 5-day notification window, respectively. Darker shading corresponds to greater percentage reductions in *R*^∗^. (c-f) The difference in the percentage reduction in *R*^∗^ that results from a 5-day and 2-day notification window, respectively, assuming that: (c) the proportion of the population who are active app users given *w* = 2 is equal to the proportion of the population who are active app users given *w* = 5, (d-f) the proportion of the population who are active app users given *w* = 5 is (d) 80%, (e) 60%, and (f) 40% of the proportion of the population who are active app users given *w* = 2.

**Figure S3:**
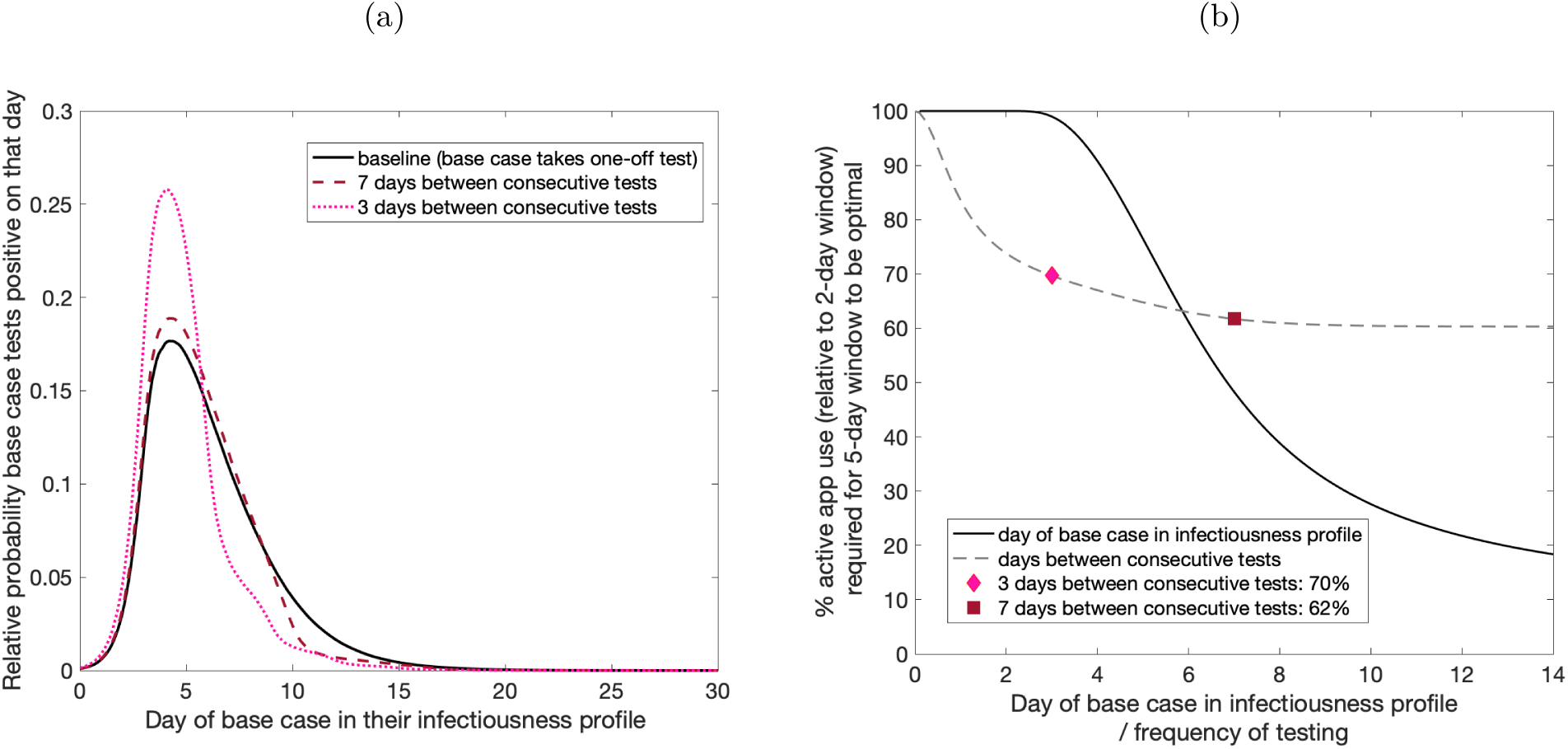
The impact of regular testing on the level of active app use required for a 5-day window to optimal notification windows: (a) The relative probability of a base case testing positive on a given day in their infectiousness profile, assuming they take a one-off test (black, solid line), assuming they take a test every seven days (dark red, dashed line), and assuming they take a test every three days (pink, dotted line). (b) The percentage active app use relative to the level of active app use given a 2-day window required for a 5-day notification window to result in a lower *R*^∗^ when: the base case is detected on day *d* (black solid line); the base case takes a test every *d* days (grey dashed line). We highlight two examples: assuming a base case takes a lateral flow test every seven days, a 5-day window is optimal given that active app use is at least 62% of the level of active app use given a 2-day window (purple square); assuming a base case takes a lateral flow test every three days, a 5-day window is optimal given that active app use is at least 70% of the level of active app use given a 2-day window (Figure S3b, pink diamond)

## Notes

### Competing Interest Statement

The authors have declared no competing interest.

### Funding Statement

TL, MJK and MJT were supported by UKRI through the JUNIPER modelling consortium [grant numberMR/V038613/1]. MJK, RNT and MJT were supported by the Engineering and Physical Sciences ResearchCouncil through the MathSys CDT [grant number EP/S022244/1]. EMH, MJK and MJT were supportedby the Biotechnology and Biological Sciences Research Council [grant number: BB/S01750X/1]. RNTwas supported by UKRI through the Rapid Assistance in Modelling the Pandemic continuity grant [grantnumber: EP/V053507/1]. MJK was supported by the National Institute for Health Research (NIHR) [PolicyResearch Programme, Mathematical & Economic Modelling for Vaccination and Immunisation Evaluation,and Emergency Response; NIHR200411]. MJK is affiliated to the National Institute for Health ResearchHealth Protection Research Unit (NIHR HPRU) in Gastrointestinal Infections at University of Liverpoolin partnership with UK Health Security Agency (UKHSA), in collaboration with University of Warwick.MJK is also affiliated to the National Institute for Health Research Health Protection Research Unit (NIHRHPRU) in Genomics and Enabling Data at University of Warwick in partnership with UK Health SecurityAgency (UKHSA). The views expressed are those of the author(s) and not necessarily those of the NHS, theNIHR, the Department of Health and Social Care or UK Health Security Agency. The funders had no rolein study design, data collection and analysis, decision to publish, or preparation of the manuscript

